# Real World Evidence of Effectiveness of COVID-19 Vaccines and Anti SARS-CoV-2 Monoclonal Antibodies Against Post-Acute Sequelae of SARS-CoV-2 Infection

**DOI:** 10.1101/2022.06.30.22277105

**Authors:** Jonika Tannous, Alan P. Pan, Thomas Potter, Abdulaziz Bako, Katharine Dlouhy, Ashley Drews, H. Dirk Sostman, Farhaan S. Vahidy

## Abstract

**Background:** We evaluated the effectiveness of COVID-19 vaccines and monoclonal antibodies (mAb) against Post-Acute Sequelae of SARS-CoV-2 infection (PASC), an emerging public health problem.

**Methods and Findings:** In a retrospective cohort study, we identified patients with clinically significant PASC using a COVID-19 specific, electronic medical record-based surveillance and outcomes registry from an 8-hospital tertiary healthcare system in the greater Houston metropolitan (primary analyses). Analyses were then replicated across a global research network database. We included all adults (>= 18) who survived beyond 28-days of their index infection. PASC was defined as experiencing constitutional (palpitations, malaise / fatigue, headache) or systemic (sleep disorder, shortness of breath, mood / anxiety disorders, cough, and cognitive impairment) symptoms beyond 28-day post-infection period. Instances of PASC were excluded if the symptoms were present pre-COVID or if they resolved within four weeks of initial infection. We fit multivariable logistic regression models and report estimated likelihood of PASC associated with vaccination or mAb treatment as adjusted odds ratios (aOR) with 95% confidence intervals (CI).

Primary analyses included 53,239 subjects (54.9% female), of whom 5,929, 11.1% (CI: 10.9 - 11.4), experienced PASC. Both, vaccinated breakthrough cases (vs. unvaccinated) and mAb treated patients (vs. untreated) had lower likelihoods for developing PASC, aOR (CI): 0.58 (0.52, 0.66), and 0.77 (0.69, 0.86), respectively. Vaccination was associated with decreased odds of developing all constitutional and systemic symptoms except for taste and smell changes. For all symptoms, vaccination was associated with lower likelihood of experiencing PASC compared to mAb treatment. Replication analysis found almost identical frequency of PASC (11.2%) and similar protective effects against PASC for the COVID-19 vaccine: aOR (CI) 0.25 (0.21 - 0.30) and mAb treatment: 0.62 (0.59 - 0.66).

**Discussion:** Although both COVID-19 vaccines and mAbs decreased the likelihood of PASC, at present, vaccination is the most effective tool to potentially prevent long-term clinical and socio-economic consequences of COVID-19.

## Introduction

Persistence or re-emergence of a constellation of symptoms and conditions weeks or months after contracting Severe Acute Respiratory Syndrome Coronavirus 2 (SARS-CoV-2) has been referred to as Post-Acute Sequelae of SARS-CoV-2 infection (PASC).^1^ The epidemiology and biology of PASC is yet to be established and there are no specific therapeutic options to ameliorate or prevent PASC. Though there is limited, and emerging evidence of COVID-19 vaccine induced protection against PASC,^2^ the real-world effectiveness (RWE) of either COVID-19 vaccination or use of Anti-SARS-CoV-2 Monoclonal Antibodies (mAbs) has not been systematically demonstrated, particularly in the United States (US). We evaluated RWE of COVID-19 vaccines and Anti-SARS-CoV-2 mAbs against PASC in a diverse US metropolitan population and further replicated our findings utilizing a global research network database.

## Methods

Our primary analyses entailed a retrospective cohort study design utilizing the Houston Methodist COVID-19 Surveillance and Outcomes Registry (CURATOR). Houston Methodist is an 8-hospital tertiary healthcare system with an extensive primary and emergency care network across the greater Houston Texas Region, serving one of the most diverse US populations of around 7 million. Detailed design and rationale of CURATOR have been previously reported.^3^ Briefly, CURATOR is an institutional review board (IRB) approved COVID-19 specific bioinformatics pipeline that captures socio-demographic, comorbidity, disease severity, hospitalization, treatment—including mAbs, and outcomes data on all COVID-19 phenotypes (tested positive / negative, hospitalized / non-hospitalized, and / or vaccinated). CURATOR is a longitudinal data repository with > 90% of patients having data on retrospective pre-COVID (since March 2016) encounters, and all patients having prospective post-COVID healthcare utilization encounters across the Houston Methodist system. Houston Methodist was one of the first state-designated vaccination centers for the greater Houston metropolitan area and initiated its vaccination campaign as per a risk-based tiered approach on December 15, 2020. Anti-SARS-CoV-2 mAbs were administered as per consistently defined clinical criteria following emergency use authorization of mAb treatment for COVID-19 by the U.S. Food and Drug Administration.^4^

Utilizing CURATOR, we identified adult patients (≥ 18 years) with a positive Polymerized Chain Reaction (PCR) test result for COVID-19 and flagged those who survived beyond 28-days of their initial diagnosis. We identified PASC based on the reported new onset of constitutional (palpitations, malaise / fatigue, and headache) or systemic (sleep disorders, shortness of breath, mood / anxiety disorders, cough, and cognitive impairment) symptoms / conditions as defined by the Centers for Disease Control and Prevention (CDC).^5^ Patients were not considered to have PASC if PASC symptoms/conditions were present pre-COVID or if they resolved within four weeks of COVID-19 diagnosis. Vaccine efficacy against PASC was evaluated among breakthrough cases only, which were defined as cases with positive PCR tests after achieving complete immunization (> 14 days after 2-doses of mRNA vaccines or a single dose of the Ad26.COV2.S vaccine).

We fit multivariable logistic regression models and report adjusted odds ratios (aOR) and 95% confidence intervals (CI) as likelihood estimates of PASC associated with COVID-19 vaccination status and mAb treatment. Models were adjusted for age (18 to 39, 40 to 64, 65+ years), sex, race, ethnicity, area deprivation index (ADI: 0 to 3, 4 to 6, 7+),^6^ Charlson Comorbidity Index (CCI: 0 to 1, 2 to 4, 5+),^7^ and COVID-19 illness severity categorized as mild/ambulatory disease (no hospitalization), hospitalized with moderate disease (with/without oxygen), and hospitalized with severe disease (intubation, ventilation, or extracorporeal membrane oxygenation).^8^

We externally validated our findings by utilizing the TriNetX Analytics Network,^9^ a de-identified global research network that comprises EMR data across 57 healthcare organizations from six different countries. Case definitions, PASC criteria, vaccination, and mAb use were similarly characterized. All analyses were replicated and adjusted for age, sex, CCI, and severe COVID-19. Other variables included in the primary analyses were missing for a significant proportion of the TriNetX sample. All analyses were conducted with the statistical software ‘R’ version 4.1.0.

## Results

In our primary analyses (March 3, 2020, to November 20, 2021), we identified 55,192 adult PCR positive patients, of whom 1,953 (3.5%) were excluded due to missing or unverifiable data. Our final cohort of 53,239 subjects (females: 54.9%, mean [sd] age: 51.6 [17.8] years, median [IQR] CCI: 1 [0, 2]) included 3,781 (7.1%) breakthrough COVID-19 cases and 4,635 (8.7%) mAb treated patients. Furthermore, 70.6% of our sample had mild ambulatory disease, 23.0% were hospitalized with moderate disease, and 6.4% were hospitalized with severe disease. Overall, 5,929, 11.1% (CI: 10.9 - 11.4), met the PASC criteria. The demographic and clinical characteristics of individuals with and without PASC are presented in Table 1.

**Table 1:**
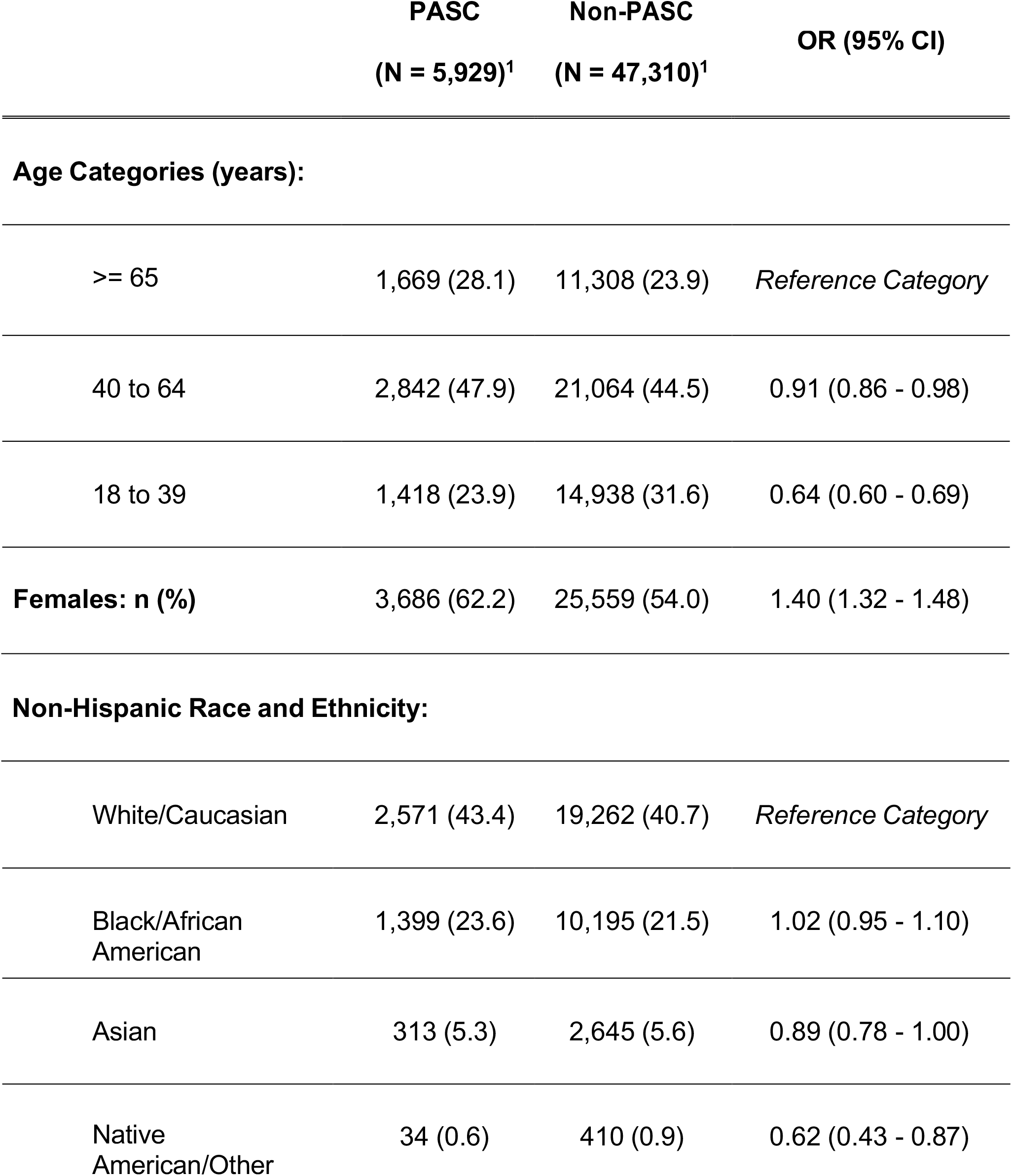
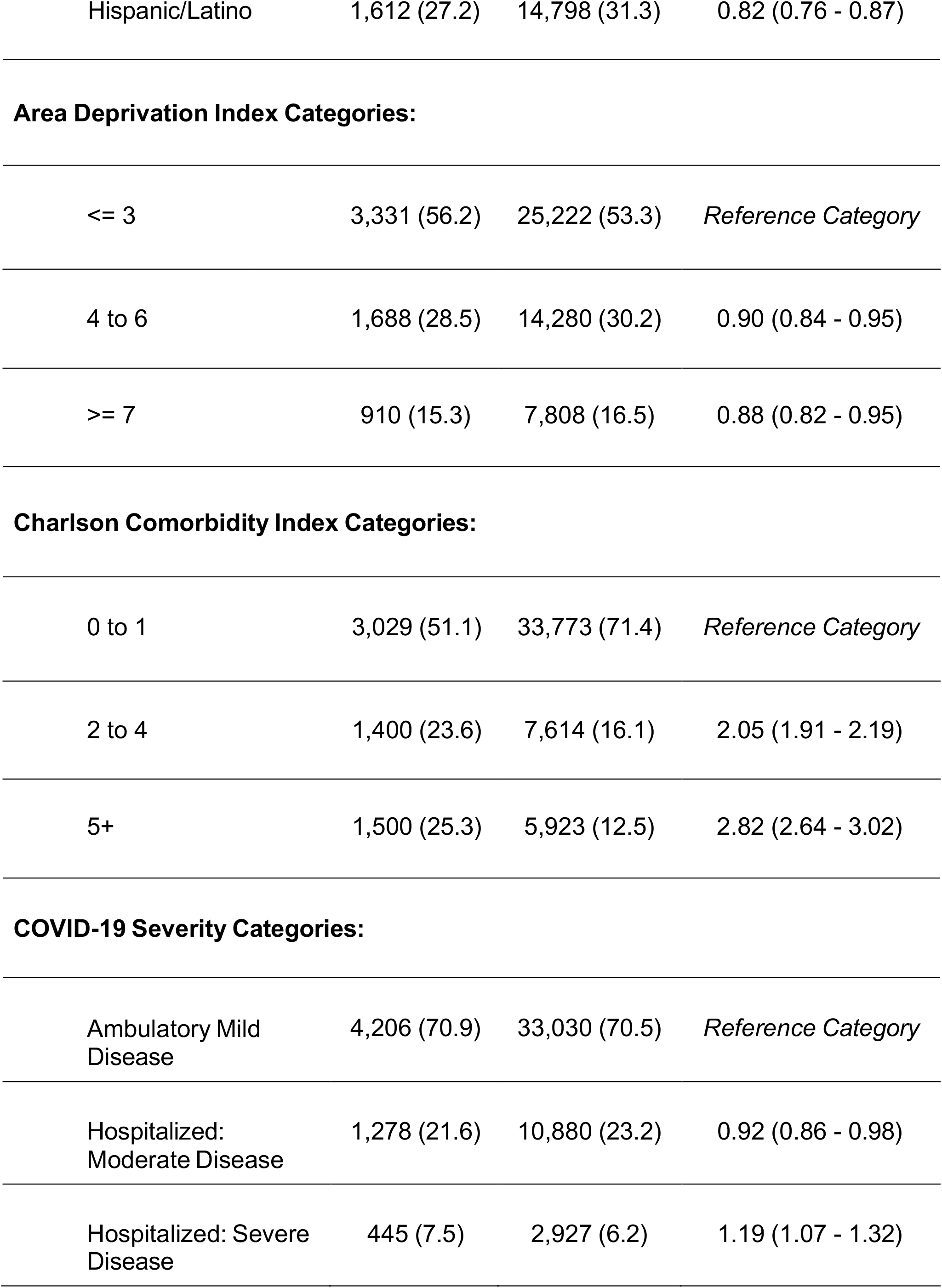
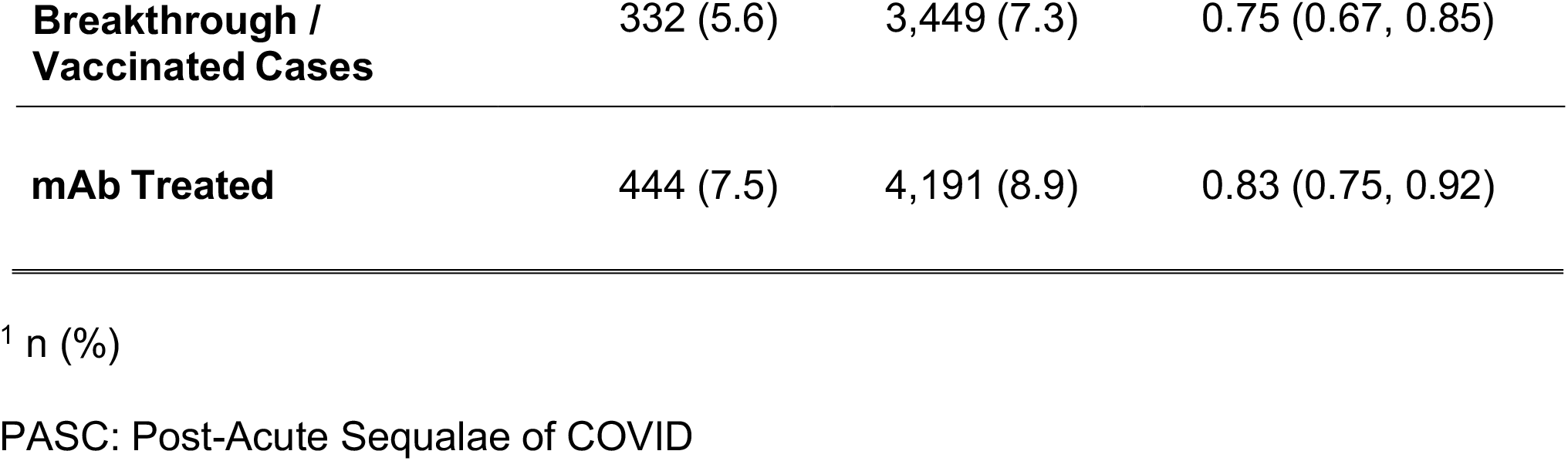
Demographic, social, comorbidity, and clinical characteristics of individuals with and without Post-Acute Sequelae of SARS-CoV-2 infection (PASC).

|  | **PASC**<br>**(N = 5,929)<sup>1</sup>** | **Non-PASC**<br>**(N = 47,310)<sup>1</sup>** | **OR (95% CI)** |
| --- | --- | --- | --- |
| **Age Categories (years):** |  |  |  |
| >= 65 | 1,669 (28.1) | 11,308 (23.9) | *Reference Category* |
| 40 to 64 | 2,842 (47.9) | 21,064 (44.5) | 0.91 (0.86 - 0.98) |
| 18 to 39 | 1,418 (23.9) | 14,938 (31.6) | 0.64 (0.60 - 0.69) |
| **Females: n (%)** | 3,686 (62.2) | 25,559 (54.0) | 1.40 (1.32 - 1.48) |
| **Non-Hispanic Race and Ethnicity:** |  |  |  |
| White/Caucasian | 2,571 (43.4) | 19,262 (40.7) | *Reference Category* |
| Black/African American | 1,399 (23.6) | 10,195 (21.5) | 1.02 (0.95 - 1.10) |
| Asian | 313 (5.3) | 2,645 (5.6) | 0.89 (0.78 - 1.00) |
| Native American/Other | 34 (0.6) | 410 (0.9) | 0.62 (0.43 - 0.87) |
| Hispanic/Latino | 1,612 (27.2) | 14,798 (31.3) | 0.82 (0.76 - 0.87) |
| Area Deprivation Index Categories: |  |  |  |
| <= 3 | 3,331 (56.2) | 25,222 (53.3) | *Reference Category* |
| 4 to 6 | 1,688 (28.5) | 14,280 (30.2) | 0.90 (0.84 - 0.95) |
| >= 7 | 910 (15.3) | 7,808 (16.5) | 0.88 (0.82 - 0.95) |
| Charlson Comorbidity Index Categories: |  |  |  |
| 0 to 1 | 3,029 (51.1) | 33,773 (71.4) | *Reference Category* |
| 2 to 4 | 1,400 (23.6) | 7,614 (16.1) | 2.05 (1.91 - 2.19) |
| 5+ | 1,500 (25.3) | 5,923 (12.5) | 2.82 (2.64 - 3.02) |
| COVID-19 Severity Categories: |  |  |  |
| Ambulatory Mild Disease | 4,206 (70.9) | 33,030 (70.5) | *Reference Category* |
| Hospitalized: Moderate Disease | 1,278 (21.6) | 10,880 (23.2) | 0.92 (0.86 - 0.98) |
| Hospitalized: Severe Disease | 445 (7.5) | 2,927 (6.2) | 1.19 (1.07 - 1.32) |
| **Breakthrough /**<br>**Vaccinated Cases** | 332 (5.6) | 3,449 (7.3) | 0.75 (0.67, 0.85) |
| **mAb Treated** | 444 (7.5) | 4,191 (8.9) | 0.83 (0.75, 0.92) |

<sup>1</sup> n (%)
PASC: Post-Acute Sequelae of COVID

In the fully adjusted models, both vaccinated (breakthrough) COVID-19 cases (vs. unvaccinated) and anti-SARS-CoV-2 mAb treated patients (vs. untreated) had a lower likelihood for developing PASC, aOR (CI): 0.58 (0.52 - 0.66), and 0.77 (0.69 - 0.86), respectively. Additionally, females (vs. males) were more likely to experience PASC [aOR (CI): 1.52 (1.44 - 1.61)], as were middle-aged (40 to 65 years) COVID-19 survivors compared to older individuals (≥ 65 years) [aOR (CI): 1.25 (1.17 - 1.34)]. Compared to COVID-19 patients with low CCI scores (0 – 1), patients with CCI of 2 to 4 and those with CCI of 5+ had the higher likelihood of experiencing PASC, aORs (CI): 2.21(2.06 - 2.38) and 3.30 (3.05 - 3.56), respectively. Individuals with higher ADI scores (vs. ADI 0 – 3) demonstrated lower odds of experiencing PASC, ADI 4 to 6: aOR (CI) 0.87 (0.81 - 0.92), 7+: 0.81 (0.74 - 0.88) and patients hospitalized with moderate disease also were less likely to have experience PASC compared to mild ambulatory cases, aOR (CI) 0.81 (0.76 - 0.87). Figure 1a provides a schematic representation of factors associated with PASC including protective effects sizes for COVID-19 vaccination and anti-SARS-CoV-2 mAbs.

**Figure 1:**
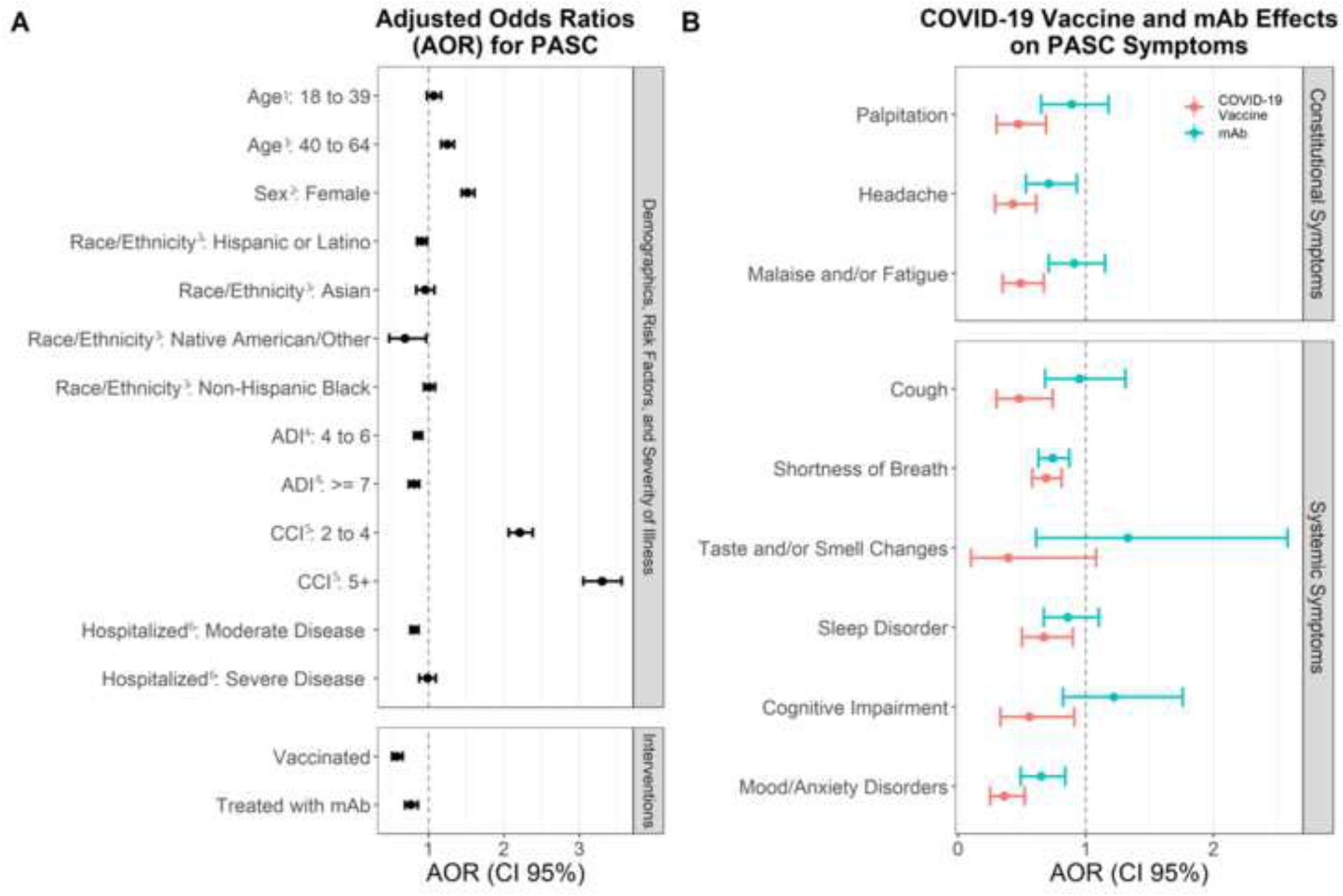
Factors associated with PASC and individual PASC symptom / conditions for breakthrough and mAb treated COVID-19 survivors. A) Likelihood of developing PASC. aORs (95% CI) are shown for vaccines and mAb along with all covariates. ^1^compared to age 65+; ^2^compared to males; ^3^compared to Non-Hispanice White/Caucasian; ^4^compared to ADI 0 to 3; ^5^compared to CCI 0 to 1; ^6^compared to ambulatory; mild COVID-19 disease. B) aORs (CI) of developing individual PASC symptoms for vaccines (red) and mAb (blue). Abbreviations: PASC: Post-Acute Sequelae of SARS-CoV-2 infection; aOR: adjusted odds ratio; CI: confidence interval; mAb: monoclonal antibodies; ADI: area deprivation index; CCI: Charlson Comorbidity Index

The frequencies of constitutional and systemic PASC symptoms among the overall, the vaccinated and the mAb treated individuals are presented in Table 2. Shortness of breath was the most common symptom, observed among 2,578 (43.5%) PASC patients. This was followed by mood/anxiety disorders, 1,001 (16.9%), and sleep disorders, 957 (16.1%). COVID-19 vaccination was associated with decreased odds of developing all constitutional and systemic symptoms except for changes in taste and smell, whereas mAb was associated with decreased likelihood of persistent shortness of breath and the new onset of mood/anxiety disorders. Across all PASC symptoms, vaccines conferred a relatively lower likelihood (greater protection) of experiencing PASC as compared to mAb treatment (Table 2).

**Table 2:** Frequencies of individual constitutional and systemic symptoms in the overall PASC group and among the vaccinated and mAb treated individuals.

|  | **Total PASC**<br>**(N = 5,929)<sub>1</sub>** | **Vaccinated**<br>**PASC**<br>**(N = 332)<sub>1</sub>** | **mAb Treated**<br>**PASC**<br>**(N = 444)<sub>1</sub>** |
| --- | --- | --- | --- |
| **Constitutional Symptoms:** |  |  |  |
| Palpitation | 591 (10.0) | 25 (7.5) | 50 (11.3) |
| Headache | 832 (14.0) | 30 (9.0) | 52 (11.7) |
| Malaise and/or<br>Fatigue | 882 (14.9) | 42 (12.7) | 76 (17.1) |
| **Systemic Symptoms:** |  |  |  |
| Cough | 474 (8.0) | 21 (6.3) | 41 (9.2) |
| Shortness of<br>Breath | 2,578 (43.5) | 160 (50.9) | 185 (41.7) |
| Taste and/or<br>Smell Changes | 74 (1.2) | 3 (0.9) | 9 (2.0) |
| Sleep Disorder | 957 (16.1) | 50 (15.1) | 71 (16.0) |
| Cognitive<br>Impairment | 248 (4.2) | 17 (5.1) | 32 (7.2) |

|  | **Total PASC**<br>**(N = 5,929)<sup>1</sup>** | **Vaccinated**<br>**PASC**<br>**(N = 332)<sup>1</sup>** | **mAb Treated**<br>**PASC**<br>**(N = 444)<sup>1</sup>** |
| --- | --- | --- | --- |
| Mood/Anxiety<br>Disorders | 1,001 (16.9) | 31 (9.3) | 59 (13.3) |

Abbreviations: PASC: Post-Acute Sequelae of SARS-CoV-2 infection.
<sup>1</sup> n (%)

The replication dataset included 631,683 patients (females: 55.4%, mean [sd] 48.7 [18.1] years. Among whom, 70,648, 11.2% (CI: 11.1 - 11.3) were identified as experiencing PASC. Similar protective effects against PASC were observed for COVID-19 vaccine: aOR (CI) 0.25 (0.21 - 0.30) and mAb treatment: 0.62 (0.59 - 0.66). Whereas female sex: aOR (CI) 1.48 (1.46 - 1.51) and higher CCI (2 to 4 vs. 0 – 1): aOR (CI) 2.13 (2.09 - 2.18) and (5+ vs. 0-1): 2.84 (2.77, 2.91) was associated with higher likelihood of PASC. Mood / anxiety disorders were the most frequently reported PASC symptom (24.4%), followed by shortness of breath (22.0%) and malaise / fatigue (21.0%). Both vaccines and mAb decreased likelihood of developing all PASC symptoms / conditions. As with our primary analysis, the likelihood of PASC reduction was consistently greater among vaccinated breakthrough COVID-19 cases as compared to these who were treated by anti-SARS-CoV-2 mAb’s. Results of the replication analysis are reported in Supplementary Tables 1-2 and Figure 1.

## Discussion

PASC has the potential to become a formidable public health challenge;^10^ and despite unresolved case-definitions and unexplained underlying pathophysiology, the phenomenon is well documented.^11^ Beyond conservative management, there are no specific treatment options.^12^

Based on our case definitions, in our primary analyses, 11.1% of COVID-19 survivors experienced PASC beyond the conventionally defined 4-week duration, which were highly corroborated (11.2%) by our replication analyses of over half a million COVID-19 patients. Prior estimates of PASC or ‘long-COVID’, utilizing direct surveys among smaller numbers of patients, primarily from European countries, vary drastically between 10% and 80%.^13^ Our literature review informs that this wide range reflects differences across studies in conditions / symptoms included in PASC criteria, the time lag between PASC assessment and acute COVID-19 illness, and perhaps most importantly the survey methodology applied. Most studies reporting higher PASC / long-COVID estimates implemented open ended questions potentially without excluding COVID-19 survivors with prior histories of similar conditions / symptoms. In fact, a prior study of 4,182 participants that prospectively assessed SARS-CoV-2 PCR positive patients for pre-specified conditions utilizing a ≥ 28-day post-COVID-19 timepoint, while excluding patients with prior history of such conditions, reported PASC estimates very similar to our analyses (13.3%, CI: 12.3 – 14.4).^14^

Given that we utilized clinical encounters for identifying PASC, it is possible that our estimates represent a more severe or clinically significant PASC experience for which medical care was sought by COVID-19 survivors. Regardless, even with potential underestimation, our data suggests approximately five million individuals across the US may be experiencing PASC, which translates into major direct and indirect healthcare and socio-economic consequences.

Our analyses uniquely demonstrate evidence of RWE of both COVID-19 vaccine and use of Anti SARS-CoV-2 mAbs against PASC. Prior reports from the United Kingdom and France have demonstrated that individuals with either completed or partial COVID-19 vaccination series are less likely to have long-term COVID-19 symptoms.^15,16^ However, to our knowledge, no direct comparisons across US populations exist. It is likely that PASC has a multifactorial etiology involving a combination of a heightened and prolonged immune response,^17^ an autoantibody driven immunomodulatory process,^18^ and even an active or persistent SARS-CoV-2 infection.^19^ Therefore, it is plausible that a boosted immunity either through vaccination or mAbs may confer protection against PASC. We found vaccination to be potentially more effective at preventing PASC than mAb. This may be explained by vaccines providing longer-term protective immunomodulation, as well as higher levels of cellular immunity and viral neutralization. Furthermore, though not causal in nature, our data seem to suggest that PASC are increasingly experienced by individuals across their prime economically productive life years. These findings corroborate with a prior report demonstrating poor post-COVID-19 recovery among those aged 40 – 59 years (compared to younger or older individuals).^20^ All together, these findings have important public health implications particularly with regards to widespread misinformation and persistent vaccine hesitancy.

Our findings also confirm the previously reported higher likelihood of PASC among females.^20^ Differential regulation of immune response between females and males is well documented,^21^ and although there is ongoing research to understand precise mechanism of PASC, the differences in underlying immune response are postulated to drive sex differences in the development of PASC. We report a relative lack of association between experiencing PASC and severity of acute COVID-19 illness. Though a prior study demonstrated lower likelihood of full post-COVID-19 recovery among patients who received mechanical ventilation,^20^ it was not ascertainable if PASC was clearly distinguishable from a broader post-ICU syndrome in these patients. Furthermore, the authors reported that several ongoing mental and physical aspects among COVID-19 survivors were unrelated to acute severity. It is therefore likely that the full gamut of factors that encompass PASC have different biological and psycho-social mechanisms that need to be fully explored in future work.

Though our findings were replicated with data from a global research network; the limitations of our primary analyses include the use of EMR from a single healthcare system. Healthcare utilization driven case definitions may underestimate the burden of PASC and may be influenced by drivers of healthcare utilization. However, our estimates are adjusted for important demographic characteristics including measures of social deprivation. Replication of our findings particularly on data derived from prospective systematic evaluation for PASC across larger and more demographically and clinically heterogenous cohorts remains necessary.

Nevertheless, our results underscore the burgeoning population health implications of PASC and highlight the critical role of vaccinations as a pivotal public health tool in potentially mitigating risk and ameliorating long-term effects of COVID-19.

## Supporting information

Supplemental Results

## Data Availability

All data produced in the present study are available upon reasonable request to the authors after necessary approvals and processes have been completed

