## Supplemental Results for "Real World Evidence of Effectiveness of COVID-19 Vaccines and Anti SARS-CoV-2 Monoclonal Antibodies Against Post-Acute Sequelae of SARS-CoV-2 Infection"

**Supplemental Table 1:** Demographic, social, comorbidity, and clinical characteristics of individuals with and without Post-Acute Sequelae of SARS-CoV-2 infection (PASC) from the TriNetX cohort.

| | **PASC (N = 70,648)**[1] | **Non-PASC (N = 561,035)**[1] | **OR (95% CI)** |
|---|---|---|---|
| **Age Categories (years):** | | | |
| >= 65 | 17,824 (25.2) | 120,577 (21.5) | *Reference Category* |
| 40 to 64 | 31,870 (45.1) | 237,924 (42.4) | 0.70 (0.69, 0.72) |
| 18 to 39 | 20,954 (29.7) | 14,938 (31.6) | 0.91 (0.89, 0.92) |
| **Females:** | 44,501 (63.0) | 202,534 (36.1) | 1.42 (1.40, 1.45) |
| **Charlson Comorbidity Index Categories:** | | | |
| 0 to 1 | 41,994 (59.4) | 431,234 (76.9) | *Reference Category* |

| | | | |
|---|---|---|---|
| 2 to 4 | 15,927 (22.5) | 79,538 (14.2) | 2.06 (2.02, 2.10) |
| 5+ | 12,727 (18.0) | 50,263 (9.0) | |
| **Severe COVID-19:** | 902 (1.3) | 5,409 (1.0) | 1.33 (1.24, 1.42) |
| **Breakthrough / Vaccinated Cases** | 121 (0.2) | 3,226 (0.6) | 0.30 (0.25, 0.35) |
| **mAb Treated** | 1,199 (1.7) | 14,462 (2.6) | 0.65 (0.61, 0.69) |

[1] n (%)

PASC: Post-Acute Sequalae of COVID

**Supplemental Table 2:** Frequencies of individual constitutional and systemic symptoms in the overall PASC group and among the vaccinated and mAb treated individuals from the TrinetX cohort.

| | Total PASC (N = 70,648)[1] | Vaccinated PASC (N = 121)[1] | mAb Treated PASC (N = 1,199)[1] |
|---|---|---|---|
| **Constitutional Symptoms:** | | | |
| Palpitation | 6,153 (8.7) | 3 (2.5) | 84 (7.0) |
| Headache | 12,472 (17.7) | 19 (15.7) | 168 (14.0) |
| Malaise and/or Fatigue | 14,847 (21.0) | 31 (25.6) | 262 (21.9) |
| **Systemic Symptoms:** | | | |
| Cough | 11,976 (17.0) | 23 (19.0) | 235 (19.6) |
| Shortness of Breath | 15,512 (22.0) | 24 (19.8) | 310 (25.9) |
| Taste and/or Smell Changes | 2,018 (2.9) | 2 (1.7) | 26 (2.2) |
| Sleep Disorder | 11,565 (16.4) | 14 (11.6) | 222 (18.5) |

| | Total PASC (N = 70,648)[1] | Vaccinated PASC (N = 121)[1] | mAb Treated PASC (N = 1,199)[1] |
|---|---|---|---|
| Cognitive Impairment | 3,535 (5.0) | 16 (13.2) | 83 (6.9) |
| Mood/Anxiety Disorders | 17,254 (24.4) | 18 (14.9) | 203 (16.9) |

Abbreviations: PASC: Post-Acute Sequelae of SARS-CoV-2 infection.
[1] n (%)

**Supplemental Figure 1:** Factors associated with PASC and individual PASC symptom / conditions for breakthrough and mAb treated COVID-19 survivors in replication analysis.

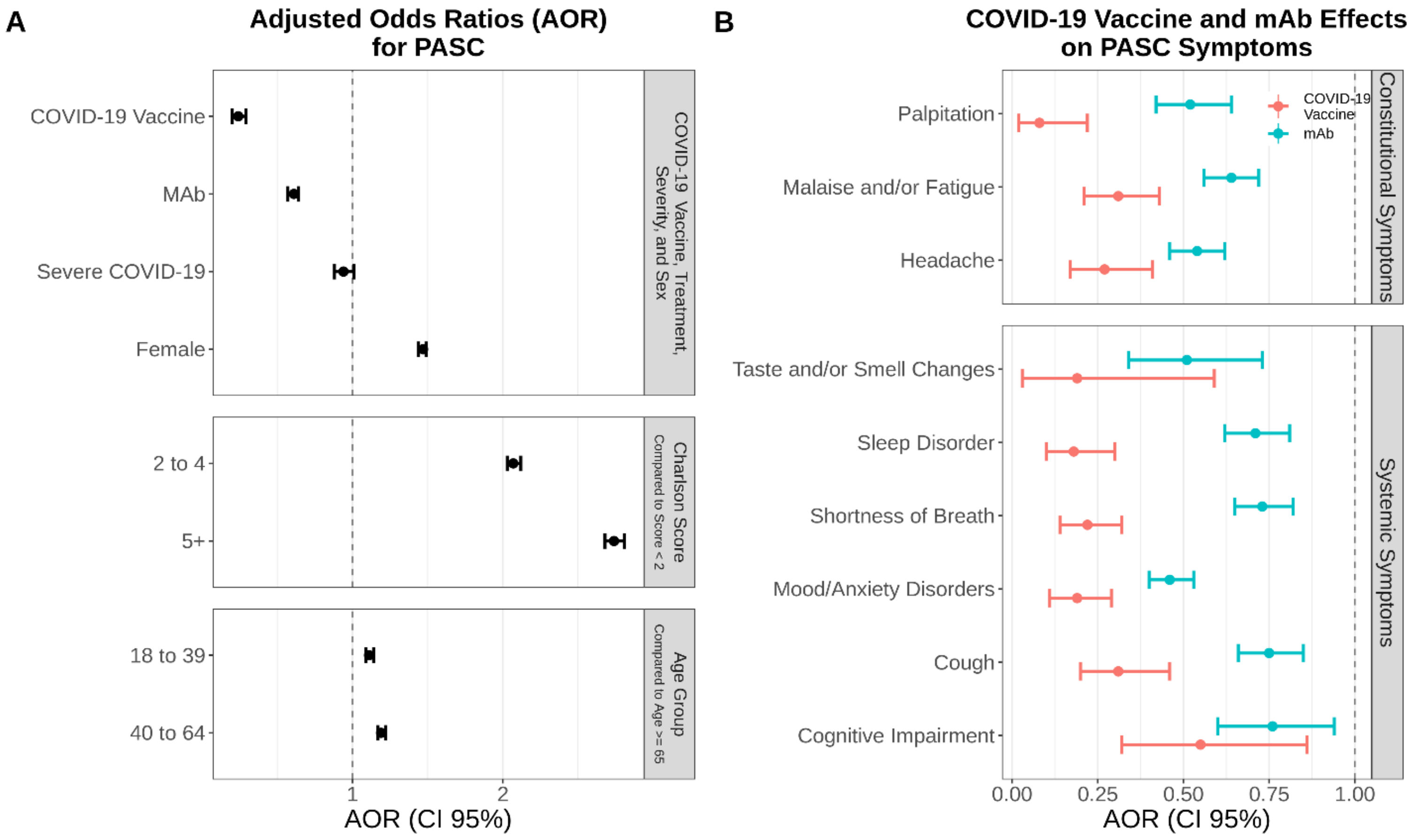


Supplemental Figure 1. A) Likelihood of developing PASC in replication analysis. aORs (95% CI) are shown for vaccines and mAb along with all covariates. B) aORs (CI) of developing individual PASC symptoms for vaccines (red) and mAb (blue) in the replication analysis.

PASC: Post-Acute Sequelae of SARS-CoV-2 infection; aOR: adjusted odds ratio; CI: confidence interval; mAb: monoclonal antibodies.
